# Measuring arterial tortuosity in the cerebrovascular system using Time-of-Flight MRI

**DOI:** 10.1101/2024.12.23.24319570

**Authors:** Yiyan Pan, Kevin Kahru, Emma Barinas-Mitchell, Tamer S. Ibrahim, Carmen Andreescu, Helmet Karim

## Abstract

The Circle of Willis (CW) is a critical cerebrovascular structure that supports collateral blood flow to maintain brain perfusion and compensate for eventual occlusions. Increased tortuosity of highrisk vessels within the CW has been implicated as a marker in the progression of cerebrovascular diseases especially in structures like the internal carotid artery (ICA). This is partly due to age-related plaque deposition or arterial stiffening. Producing reliable tortuosity measurements for vessels segmented from magnetic resonance (MR) time-of-flight (TOF) images requires precise curvature estimation, but existent methods struggle with noisy or sparse segmentation data. We introduce an open-source, end-to-end pipeline that uses unit-speed spline fitting for accurate curvature estimation, generating robust curvature-based tortuosity metrics for the ICA combined with an indicator of spline fit quality. We test this with theoretical data and apply this method to TOF data from 22 participants. We report that our metrics are able to capture tortuosity even under heightened noise constraints and discriminate different types of abnormal arterial coiling. We found that our ICA tortuosity measures correlate positively with age and ultrasound measured carotid artery intima media thickness. This ultimately has important translational implications for being able to reliably generate TOF tortuosity measures and estimate cerebrovascular disease burden. We provide open-source code in a GitHub repository.

## Introduction

The Circle of Willis (CW) is a continuous arterial circle at the brain’s base, formed by interconnections between branches of the left and right internal carotid arteries (ICA) and the vertebrobasilar system (Vrselja et al., 2014). This structure supports collateral circulation by redirecting blood flow in cases of obstruction such as ischemia or stenosis. Evolutionarily, the CW’s presence in many non-human species suggests an advantage in compensating for blood flow loss due to occlusion.

Recent studies suggest that the geometric features of specific CW arteries may serve as important markers for cerebrovascular pathologies (Hoksbergen et al., 2003). Anatomical abnormalities frequently occur in the internal carotid artery (ICA) (Raidah et al., 2023), often presenting as coiling, looping, or bending. Elevated cerebral vessel tortuosity is often a result of vascular remodeling triggered by micro infarcts from atherosclerosis and the thickening of the blood vessel walls. Highly tortuous arteries are both a biological marker of cerebrovascular damage and a risk factor for further complications as vessel tortuosity disrupts laminar blood flow, elevating the risk of plaque burden, clots and infarcts which compounds the risk of cerebrovascular accidents (CVAs).

Altered cerebral blood flow is a leading cause of longterm disability, and CVAs are the third leading cause of death (Naghavi et al., 2024). Importantly, there is an asymptomatic period, also called “silent interval”, between the onset of changes in vessel tortuosity and the manifestation of CVAs clinical symptoms. During this silent asymptomatic period, vessel tortuosity increases as an adaptive response to alterations in blood flow or pressure(Saba et al., 2021). Being able to quantify arterial tortuosity during this preclinical phase offers insights into the disease progression and can serve as an early biomarker of potential cerebrovascular complications, aiding in timely intervention and management.

A major challenge in using the full geometric profile of all the CW arteries as a marker is that intracerebral structures are inherently tortuous with significant individual variation. This complicates the detection process as individuals’ CW may deviate from a single, standardized shape for intracerebral arteries (Bullitt et al., 2003). Assessing the geometry through tortuosity of isolated arteries such as the ICA—where most abnormalities occur (Raidah et al., 2023)—is a more practical and targeted approach.

Despite recent advancement in fully automated, end-to-end methods which produce accurate segmentation of CW arteries with anatomical labels from TOF images (Dumais et al., 2022), there are no open source tools for extracting evaluation of tortuosity based on common tortuosity metrics like Arc over Chord (also called Tortuosity Index), Total Curvature, Mean Squared Curvature, and Root Mean Squared Curvature. Most of these have been previously used in the literature for defining tortuosity. While the ICA typically exhibits S or U-shaped anatomical profile, it can also take on more tortuous shape characterized by higher frequency changes rather than large amplitude changes. As a result, the traditional Arc/Chord method alone may not adequately capture these forms of tortuosity. Given that the rest of the aforementioned tortuosity measurements are curvature-based (Kashyap et al., 2023), it is essential to obtain accurate curvature estimates. This is challenging because segmentation represents a discrete set of points, while curvature is defined for continuous functions. Previous research has shown that methods based on discrete sampling are highly sensitive to noise, particularly in calculating second derivatives (Johnson and Dougherty, 2007).

In this paper, we use polynomial spline fitting as proposed by Johnson et al.(Johnson and Dougherty, 2007), as direct sampling is too prone to noise. We propose that fitting a uniform spline, in conjunction with curvature-derived metrics, will yield accurate curvature estimates relevant for research applications and improves the rigor of derived measures. We create an end-to-end workflow with important quality assurance measures to compute multiple measures of tortuosity. We additionally test whether our curvature metrics correlate with age as well as maximum carotid intima-medial thickness, a surrogate marker of atherosclerosis.

## Method

### Participants and study design

The participants data was collected from an an administrative supplement to R01 MH108509 (Functional neuroanatomy correlates of worry in older adults), aiming to collect preliminary data on the risk of older severe worriers to develop Alzheimer’s disease (AD) or AD-related dementias (ADRD) (3R01MH108509-05S1). We recruited participants (*n* = 22) who were between 50-60 years with varying levels of worry and anxiety.

Participants were recruited from the Pittsburgh area via Pitt+Me (website resource from the university), inperson recommendations, flyers, and radio/television advertisements. Exclusion criteria included uncorrected vision problems, below 6th grade reading level, MRI contraindications, autism spectrum disorder, intellectual development disorder, major neurocognitive disorder, psychosis, bipolar disorder, history of cerebrovascular accident, multiple sclerosis, vasculitis, significant head trauma, presence of Axis II disorders, increased suicide risk, drug/alcohol abuse within the last six months, uses of high doses of benzodiazepines (equivalent to >2 mg of lorazepam), or current use of antidepressants. Participants with mild cognitive impairment (MCI) were allowed in the study. This study was approved by the University of Pittsburgh Institutional Review Board. All participants gave written informed consent before participating in the study.

### Assessments

Along with demographic information (age, sex, race, and education), we acquired carotid ultrasound measurements, including intima-media thickness. We used an Acuson Sonoline Antares (Siemens) high resolution duplex ultrasound scanner to obtain bilateral carotid images of the common carotid artery (CCA), carotid bulb and ICA. Digitized images obtained at end-diastole of these segments were read using semi-automated software to measure IMT by electronically tracing the lumen-intima interface and the media-adventitia interface at the following 8 locations (4 from left and right carotid arteries): near and far walls of the distal CCA, and far walls of the carotid bulb and ICA. The computer then generates one measurement for each pixel over these areas, and the maximum of all average readings across the eight locations comprised the average carotid IMT. We call this measure Mmax. We also collected data on worry (Penn-State Worry Questionnaire, PSWQ) (Meyer et al., 1990), anxiety (Hamilton Anxiety Rating Scale, HARS) (Hamilton, 1959), and measures of systolic and diastolic blood pressure (BP).

### MRI Data Acquisition

MR data were acquired on a 7T Siemens Magnetom scanner on a custom 16-transmit and 32-receive channels (Tic-Tac-Toe design) radiofrequency (RF) coil system (Santini et al., 2020) operating in the single transmit mode (Santini et al., 2018; Krishnamurthy et al., 2019). Structural images were acquired using a 3D multiecho Magnetization Prepared Rapid Gradient Echo (MPRAGE) sequence (TR = 3000ms, TE = 2.17 ms, TI = 1200 ms, 0.75 mm^3^ isotropic resolution, and GRAPPA acceleration factor of 2).). TOF images covering the whole brain were acquired using TOF 7 slabs protocol with with TR: 14 ms, TE 4.5 ms, resolution = 0.38 mm^3^, and 17.86 mm slab overlap.

### Vessel Segmentation

We use the open-source Express Intracranial Arteries Breakdown (eICAB) method developed by Dumais et al. (Dumais et al., 2022) for automated segmentation and labeling of the CW arteries from input TOF images. The pipeline generates an image annotated with 18 labels corresponding to major arteries, with a resolution of 0.625 mm^3^ isotropic. The segmented output is aligned in the same spatial orientation as the resampled TOF. To enhance the segmentations quality, manual corrections were performed using ITK-SNAP (Yushkevich et al., 2006) addressing disconnected regions of the same structure. Supplementary Figure S1 illustrates examples where manual adjustments were applied (indicated with arrowhead) on the segmentations of the left (green label) and right (red label) ICA.

### Data Description

After running the eICAB pipeline, we retained only highquality segmentation (*n* = 11) from the PALS dataset, which serves as our evaluation set for the developed metrics. The dataset has a mean age of 56.6 years ±3.2, with 3 males and 8 females. The mean Mmax value is 0.904 ± 0.080. For the psychological scores, the mean PSWQ Total is 53.8 ± 12.3, the mean HARS score is 14.7 ± 8.3, the mean systolic blood pressure(BP) is 127.4 ± 15.7, and the mean diastolic BP is 80.6 ± 7.3 (Table 1).

**Table 1.** Demographic of PALS Dataset.

|  | Mean | Standard Deviation |
| --- | --- | --- |
| **Age** | 56.6 | 3.2 |
| **Mmax** | 0.904 | 0.080 |
| **PSWQ Total** | 53.8 | 12.3 |
| **HARS Score** | 14.7 | 8.3 |
| **Systolic BP** | 127.4 | 15.7 |
| **Diastolic BP** | 80.6 | 7.3 |

### Tortuosity Metrics and Curvature Estimation

Estimating the tortuosity of individual large arteries is important when evaluating atherosclerosis since abnormal tortuosity is associated with both increased risk of stroke and failure of endovascular therapy (Bullitt et al., 2003). The most common vascular tortuosity metrics are Arc over Chord, which measure the actual path length over the Euclidean distance between two endpoints of a vessel. This metric is extremely popular due to its ease of calculation, but it has the disadvantage of being insensitive to arteries that “wiggle” (Capowski et al., 1995). Given that the skeleton vessel could be represented as a discrete space curve in 3D space, and Curvature and Torsion are the most important properties for describing how curves bend in 3D (Nguyen and Debled-Rennesson, 2008), we included the most commonly used curvature-based tortuosity metrics including Mean Curvature, Mean Squared Curvature, and Root Mean Squared Curvature in (Table S1).

The Mean (Total) Curvature *κ*_*m*_ is the integral of curvature along the length of the artery and is scale invariant. The Mean Squared Curvature is similar to *κ*_*m*_ but applies higher weights to greater curvature values in the summation. The normalized root mean squared curvature *κ*_*rms*_ is similar to *κ*_*ms*_ but because of the normalization with arc-length, this metric is scale invariant like *κ*_*m*_.

Given that 3 out of 4 popular tortuosity metrics we are referencing are curvature-based, the first step to output metrics is to have accurate approximation method to acquire the curvature estimation from the space curve of the segmented arteries. Popular approximation methods of curvature includes estimating the derivatives of the curve using finite difference. Velocity and Acceleration vectors are calculated from consecutive discrete points(Bullitt et al., 2003). Frenet-Seret frame (comprising the Tangent, Normal, and Binormal vectors) is utilized to compute curvature based on the rate of change of the tangent vector along the curve. This method is inherently local and its approximations can be sensitive to noise, leading to significant errors in curvature estimation if the data is noisy. Circle fitting could approximate the curvature at a point on the curve by fitting a local circle (osculating circle) to a neighborhood of points around that point(Kashyap et al., 2023). The inverse of the circle’s radius is then used as an estimate of the curvature. Circle fitting is moderately scalable for larger datasets, but it is limited to regions where circular approximation is valid and requires careful selection of sliding window size. Spline fitting interpolates the discrete points of the space curve with a smooth parametric spline (e.g., cubic spline, B-spline). Once the spline is fit, curvature can be analytically derived from the splines’ parametric equations using derivatives. Spline fitting is computationally expensive and does not scale well with large datasets but it handles varying curvature smoothly.

Given the noise present in our TOF in addition to the noise introduced by segmentation, the direct numerical approximation method does not perform well especially when the points that constitutes the space curve of arteries have varied distance between them. The circle fitting also does not perform well as one tortuous space curve has varied curvature along its length, and will require us to get an optimal window of points. This is difficult to do especially when the skeleton of artery segmentation output only consists of on average of less than 30 points. After testing all 3 approximations, we believe fitting unit-speed spline to the artery mid-line skeleton is the best way to approximate curvature for space curve of artery as it is more robust to noise than finite difference direct approximation, and it avoids manually choosing a window size or fitting on a limited number of points for circle fitting.

### Work Flow from Segmentation to Tortuosity Metrics

To quantify tortuosity from segmentation data, we build a pipeline encompassing skeletonization, endpoint detection, path ordering, spline fitting, and curvature-based tortuosity estimation. We first convert segmentations into binary masks by isolating the target label of interest (the ICA). This binarization ensures that only the anatomical structure of interest was retained for subsequent analysis. The binary 3D arrays are skeletonized using the skeletonize function from the skimage library. The skeletonization is medial-axis extracted from the cloud of points representing the segmented structure. It preserves the topological integrity of the cloud of points while reducing its representation to a single-pixel-wide line.

To identify the endpoints of each skeletal structure, we convolve each point in the skeleton with a 3×3×3 neighborhood kernel. Points with only one active neighboring voxel were marked as the terminal of the skeletal pathway. We do not consider the scenario where there are multiple end points for a single structure or bifurcation points since elCAB has already effectively handled bifurcation by assigning each branch with different labels. The convolution-based start-tail points detection is necessary for setting starting and end points. This traversal from the identified start point to the end point produces an ordered list of 3D points that accurately represents the morphological path of the structure since coherent, ordered sequence of points are a requirement for spline fitting.

Next, a uniform-speed 3rd-degree B-spline was fit to the ordered list of skeleton points. This spline provides a smooth and continuous parametric curve approximating the central trajectory of the segmented structure. The uniform-speed property of the B-spline ensures consistent parametrization along the curve, which is essential for accurate derivative computations. By computing the first and second derivatives of the B-spline, we quantify the curvature along the pathway. Higher curvature values corresponded to more tortuous segments, allowing for a precise quantitative assessment of the structural complexity. Then, the tortuosity metric can be derived from the curvature of the fitted spline as illustrated in (Figure 1). In the output CSV file, we also produce both the Root Mean Square Curvature and the Root Mean Square Error (RMSE) from the derived spline to the input points as a quality measure of our fitting. Smaller RMS Curvature correspond to smoother spline and smaller RMSE correspond to spline that has higher fidelity to the points.

**Figure 1.**
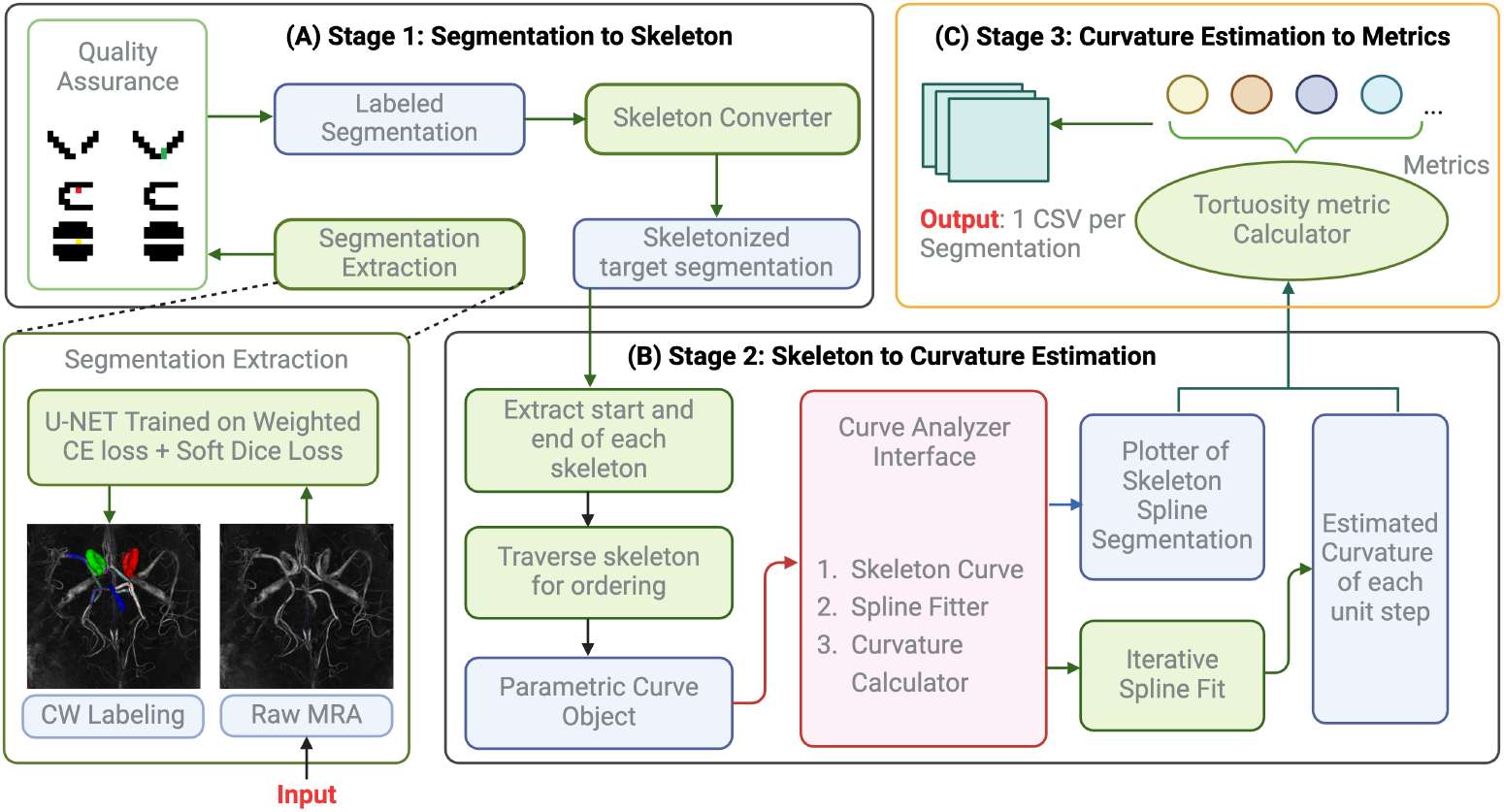
workflow of segmentation to csv file for the spline fit tortuosity metrics estimation pipeline

## Results

After building the end-to-end pipeline, it is essential to evaluate the utility of these metrics. This evaluation is conducted using both simulated parametric curves and data from the PALS study. On parametric curves like helix of varied radius and pitch, we tested how each metrics changes with radius and pitch of helix. On clinical data, we test each metric’s Spearman correlation with MMax and age.

### Phantom Helix Result

By testing on phantom helix data with same radius and different pitches, we observe that all *κ*_*m*_, *κ*_*ms*_, *κ*_*rms*_ decreases with increasing pitch as expected. This trend aligns with our intuition, as increasing the pitch while keeping the radius and number of turns fixed makes the curve progressively “straighter.” By fixing the radius and pitch and varying the number of turns, we find that all our metrics assign higher values to more tightly coiled shapes (i.e., helices with more turns while other parameters remained constant). Scale invariance was verified in *κ*_*m*_ and *κ*_*rms*_. Specifically, for helices where the radius and pitch are doubled while the number of turns remains the same, the values of *κ*_*m*_ and *κ*_*rms*_ remain unchanged, as shown in (Figure 2).

**Figure 2.**
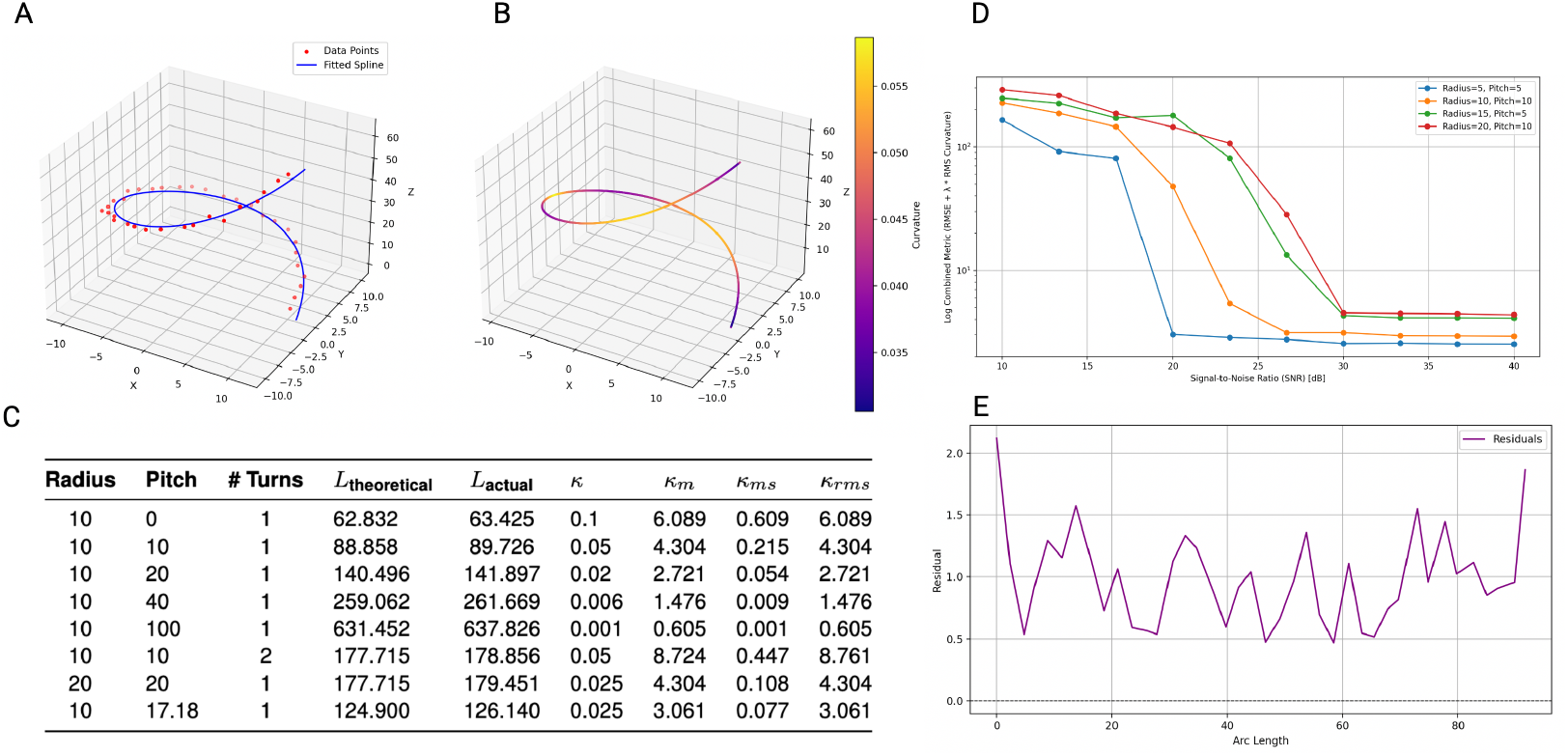
Phantom Helix Testing Result. **A**. spline fitting on helix **B**. Same spline as in A, spline colored by curvature at each point. **C**. Table of Helix with varies radius, pitch and turns and their tortuosity metrics derived from fitted spline. **D**. log scale combined metrics of rmse + rms curvature of fitted spline decrease as we increase signal to noise ratio in all multiple helices. **E**. example of residual from data point to fitted spline from helix in A, plotted along the length of this curve

### Tortuosity Correlation with PALS Age-related Variables

Computing the tortuosity metrics from our PALS data and calculating the spearman correlation of each of the metrics with *M*_*max*_ show that all curvature-based metrics are positively correlated with *M*_*max*_ yet AOC is not. (see Table 2). Similarly, all curvature-based metrics are positively correlated with age whereas AOC shows a negative correlation. (see Table 3). All Spearman’s *ρ* values were not statistically significant; even age itself has non-significant Spearman’s rho of 0.014 with Mmax and corresponding p-value of 0.968. Pairwise correlation between metrics pair shows AOC exhibits only a slight positive correlation with the curvature-based metrics. In contrast curvature-based metrics are strongly and significantly positively correlated with each other. For instance, total curvature and root mean square curvature have Spearman’s *ρ* of 0.951 with *p* value of 1.08 × 10^−11^. (see Table 4).

**Table 2.** Spearman Correlation Results for Curvature Measures vs. Mmax.

| Curvature Measure | Spearman $\rho$ | p-value |
| --- | --- | --- |
| Total Curvature | 0.039 | 0.865 |
| Mean Squared Curvature | 0.009 | 0.968 |
| RMS Curvature | 0.032 | 0.889 |
| AOC | -0.131 | 0.560 |

**Table 3.** Spearman Correlation Results for Curvature Measures vs. Age.

| Curvature Measure | Spearman $\rho$ | p-value |
| --- | --- | --- |
| Total Curvature | 0.152 | 0.500 |
| Mean Squared Curvature | 0.249 | 0.264 |
| RMS Curvature | 0.230 | 0.302 |
| AOC | -0.076 | 0.735 |

**Table 4.**
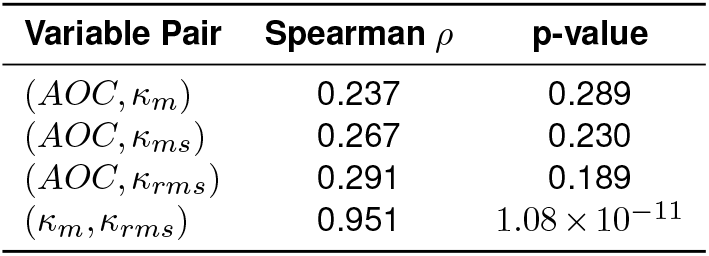
Additional Correlations.

| Variable Pair | Spearman $\rho$ | p-value |
| --- | --- | --- |
| (AOC, $\kappa_m$) | 0.237 | 0.289 |
| (AOC, $\kappa_{ms}$) | 0.267 | 0.230 |
| (AOC, $\kappa_{rms}$) | 0.291 | 0.189 |
| ($\kappa_m$, $\kappa_{rms}$) | 0.951 | $1.08 \times 10^{-11}$ |

## Discussion

This paper introduces a fully open-source, end-to-end pipeline that takes segmented TOF images as input and outputs a CSV file of tortuosity metrics. The pipeline evaluates vessel tortuosity by fitting splines optimized through minimizing root mean square error (RMSE) of curvature. Unlike previous methods that rely on non-spline-based curvature approximations, this approach is more robust to noise. Additionally, unlike other spline-fitting methods, our approach includes RMSE as a reliability indicator in the output, providing a quantitative measure of the fit’s reliability. The validity of tortuosity derived from spline fit approximation is substantiated by simulated helix data as our metrics are able to assign higher value to helices that are more tightly coiled.

Curvature-based tortuosity metrics and age both demonstrate a similar association with Mmax. IMT is a risk factor and surrogate marker for atherosclerosis, a condition that typically progresses with age (Bonithon-Kopp et al., 1996; Wang and Bennett, 2012). IMT is positively associated with the increased atherosclerotic plaque development that could lead to increased tortuosity of carotid vessel and higher blood pressure (Tschiderer et al., 2023). However, the mean systolic blood pressure of our sample at was baseline is 127.39 mmHg (SD = 14.87), and the mean diastolic blood pressure was 80.56 mmHg (SD = 7.01). Given the relatively young age range of our participants (50-60 years), these values do not indicate hypertensive levels. This suggests our PALS cohort is less likely to have the degree of arterial tortuosity observed in older(70-80 years) or more hypertensive populations, where vascular changes are typically more pronounced(Sun et al., 2022). As such, IMT did not show a statistically significant association with vessel tortuosity in our PALS dataset. We plan to validate our approach on on larger and older cohorts.

This pipeline facilitates easy calculation for tortuosity measures in CW, providing an end-to-end solution with measures that are less susceptible to noise, which allows for estimation of subthreshold changes in tortuosity, particularly during the silent interval. Ultimately, it enables researchers to understand how different clinical variables in the elderly (e.g., anxiety, worry) are associated with tortuosity and their potential association with increased risk of early cognitive decline (Zlokovic et al., 2020).

## Data Availability

All data produced in the present study are available upon reasonable request to the authors

## Appendix

Helix is generating by finding pair of radius *r* and pitch *c*

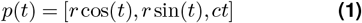

The arc length of curve *f* starting from point *t*_0_ is defined as

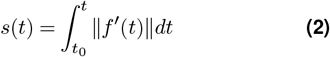

∥ · ∥ denote the Euclidean vector norm.

A curve is parametrized by arc length if t is the arc length of *f* measured from some start point such that *ds/dt* = ∥*f* ^*′*^(*t*)∥ = 1 and

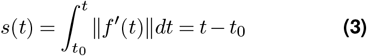

To achieve arc length parametrization numerically, the following steps were performed:

1. Fit cubic splines to the *x, y*, and *z* coordinates of the input curve *f* (*t*).
2. Compute the derivatives of the spline, *f* ^*′*^(*t*), and calculate the arc length differentials *ds* = ∥*f* ^*′*^(*t*)∥ *dt*.
3. Generate a cumulative mapping from parameter *u* to arc length *s*, and invert it to map arc length *s* back to *u*.
4. Evaluate the spline at uniformly spaced arc length values, producing a re-parametrized curve *f* (*s*).

## Supplementary Information

**Figure S1.**
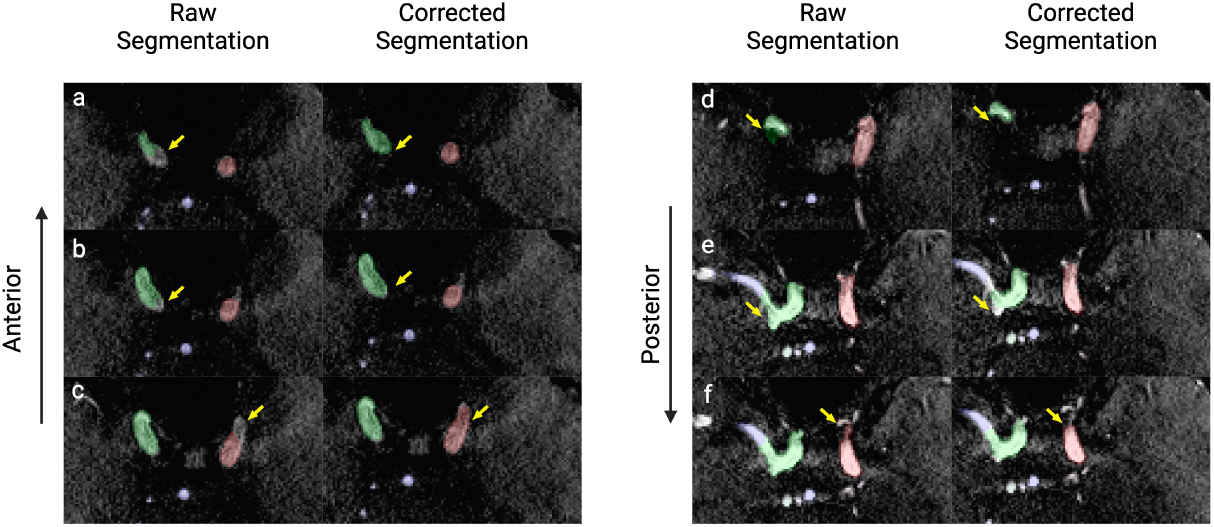
Quality Assurance example of eICAB segmentation

## Supplementary Tables

**Table S1.** Curvature and Tortuosity Metrics.

| Metric Name | Metric Symbol | Metric Formula |
| --- | --- | --- |
| Arc over Chord (AOC) | $\tau$ | $\tau = \frac{L}{C}$ |
| Mean(Total) Curvature | $\kappa_m$ | $\kappa_m = \int_{t_1}^{t_2} \kappa(t) dt$ |
| Mean(Total) Squared Curvature | $\kappa_{ms}$ | $\kappa_{ms} = \int_{t_1}^{t_2} \kappa^2(t) dt$ |
| Normalized Root Mean Squared Curvature | $\kappa_{rms}$ | $\kappa_{rms} = \sqrt{\frac{1}{L} \int_{t_1}^{t_2} \kappa^2(t) dt}$ |

## Notes

### Competing Interest Statement

The authors have declared no competing interest.

### Funding Statement

This study was funded by NIH with project name Functional Neuroanatomy Correlates of Worry in Older Adults and project number 3R01MH108509-05S1

### Author Declarations

Ethics committee/IRB of University of Pittsburgh gave ethical approval for this work

